# Evaluation of person-centred outcome measures for use in clinical trials of tuberculosis therapeutics

**DOI:** 10.1101/2025.04.07.25325380

**Authors:** Conor D Tweed, Annabelle South, Suzanne Staples, Bazarragchaa Tsogt, Nestani Tukvadze, Rebecca M Turner, Stephan Dressler, Adam T Gray, Blessina Kumar, Oxana Rucsineanu, Paul Sommerfeld, Rochelle Burgess, Hanif Esmail

## Abstract

**Background:** There is a need for person-centred outcome measures (PCOMs) in tuberculosis (TB) clinical trials to capture the priorities of affected individuals relating to both disease and treatment, alongside the traditional efficacy and safety analysis.

**Methods:** We conducted a literature search to identify existing health-related quality-of-life (HR-QoL) measurement tools and instances where they have been used to evaluate TB disease in interventional studies. DR-TB survivors were recruited for focus group discussions in South Africa, Mongolia and Georgia to determine the most important aspects of disease and treatment experience from their perspective. A second round of focus groups then worked to prioritise these items and group them within different domains. A panel of researchers and academic clinicians evaluated existing PCOMs identified in the literature search using a published framework. Finally, an assessment was carried out by a experienced TB Community Advisory Group members for appropriateness and acceptability, and clinical trial site investigators for feasibility of the PCOMs.

**Results:** The initial round of 9 focus groups involving 57 participants identified priority items grouped under the fields “overall quality of life”, “disease symptoms”, and “treatment and side effects”. In the second round of four focus groups involving 26 participants, the highest ranked items under “overall quality of life” were duration of treatment, experience of swallowing pills, and how long before being able to contribute to the household. The most important symptoms were coughing, weight loss and breathlessness. The highest ranked side effects were nausea/vomiting, skin colour changes, and nerve problems. FACIT-TB was the only measurement tool assessed that was rated “good” for appropriateness, and “excellent”, “good” or “fair” in all other fields by community representatives, researchers and clinical trial site staff.

**Conclusions:** The FACIT-TB measurement tool was identified as the most appropriate disease-specific PCOM currently available for use in clinical trials investigating novel DR-TB therapeutics.

## Introduction

The tuberculosis (TB) pandemic claims approximately 1.6 million lives per year globally. It is estimated there were over 10 million new cases of active disease in 2023, and approximately 500,000 of these were cases of drug-resistant TB (DR-TB).^1^ Traditionally, TB treatment itself is lengthy and toxic, with DR-TB treatment less effective than first-line treatment for drug-susceptible TB (DS-TB).^2^ This has a significant impact on acceptability and adherence in real-world settings and consequently on cure rates.^1^

For the first time in decades, multiple new drugs and regimens are being investigated for the treatment of both DS-TB and DR-TB in Phase 3 trials. Several of these novel regimens have already demonstrated 80 - 90% efficacy in these trials.^3-6^ However, there could be important differences relating to the lived experience of treatment and disease that are not captured through traditional efficacy and safety endpoints in trials. These differences could have a significant impact on the real-world effectiveness of these regimens by influencing adherence and also make different regimens more suitable for some people with TB over others. The GRADE (Grading of Recommendations Assessment, Development and Evaluation) Working Group has developed Evidence to Decision (EtD) frameworks to promote a structured, reproducible approach to the evaluation and implementation of evidence in guidelines.^7^ These frameworks include criteria relating to the acceptability, appropriateness and undesirable effects of interventions which would generally require more than a standard efficacy and safety analysis to determine. Additionally, the World Health Organisation (WHO) has also recently published a Guideline on Evidence Generation for New Regimens for Tuberculosis Treatment^8^ that includes an EtD framework with similar categories, which confirms the need for TB-specific PCOMs to inform guideline development.

Person-reported outcome measures (PROMs) and person-centred outcomes (PCOMs) help with evaluating a novel intervention’s real-world effectiveness by accounting for aspects of the intervention not captured by standard efficacy and safety reporting. Previous TB studies have used widely-accepted health-related quality-of-life (HR-QoL) assessment tools, such as EQ-5D or SF-36.^9-11^ However, there has been a limited amount of work done to evaluate the performance of different HR-QoL assessment tools in the context of interventional trials in TB disease and how best to capture disease-specific priorities of people with TB.^12^ The aspects of treatment, side effects, and the disease itself that matter the most to people with TB have been explored,^13^ but how best to capture these items using a clinical trial outcome measure was not investigated as part of this work.

This work sought to identify the most important aspects of treatment and disease experience according to focus groups composed of DR-TB survivors, before ranking them and selecting HR-QoL assessment tools that best capture these priorities. Our aim was to identify PCOMs for use in TB trials that could be used to capture people’s priorities relating to both disease symptoms and treatment effects.

## Methods

We applied a sequential approach to collect existing QoL measurement tools, identify the priorities of people with TB, rank these priorities, and then determine which of the existing QoL tools captured these top priorities most effectively.

### Literature Search

We conducted a literature search between January 2021 and March 2025 using PubMed, OVID, and EMBASE to identify existing HR-QoL measurement tools and instances where they have been used to evaluate TB disease in interventional studies. The search terms used included “tuberculosis” OR “TB” AND “quality of life” OR “symptoms” AND “patient-centred” AND “outcome measures”.

A review of paper titles and abstracts was conducted by CDT & AG, and quality of life scores that had been used in studies relating to TB were identified, including two systematic reviews.^12^ ^14^ These measurement tools were collated and characteristics were recorded including domains covered, items addressed, and scoring system (numerical, Likert etc).

### First Round of Focus Groups

An initial round of focus groups was carried out at sites in South Africa, Mongolia and Georgia to identify the most important themes relating to DR-TB treatment from the perspective of the person with TB. Persons aged 18 years or older who had either completed treatment for DR-TB in the previous 12 months or were in the “continuation phase” of treatment (or final 3 months if receiving a 6-month regimen) were included. Participants were clinically well and sputum culture negative. Those who were symptomatic, unwell, or sputum culture positive were excluded. The methods for this first round of focus groups have been described in more detail in an associated publication.^13^

### Second Round of Focus Groups

A second round of focus group work involving different participants was conducted at the sites in South Africa and Mongolia. This was to i) confirm that all key issues relating to treatment and disease symptoms in DR-TB had been captured in the first round before ii) ranking them according to importance for the participants.

The participants were initially presented with a list of issues identified in the first round of focus groups. A single set of cards with issues identified in the previous round of focus groups relating to treatment side effects, disease symptoms and overall quality of life were used. Participants were then asked to suggest additional issues that they believed should be included within these categories as more cards. A ranking exercise was undertaken with participants attaching stickers to cards to indicate their importance (greater number of stickers indicating greater importance) in each of the three categories. Participants were given 15 stickers each per category, and permitted to add as many of these stickers as they wished to each card with the total number of stickers on each card counted at the end. This approach was adopted as a simple, reproducible method that could be easily used by participants from a range of backgrounds.

Focus group transcripts and results of the ranking exercises were reviewed for both sites. The total number of stickers was obtained for each issue in the three categories and an overall ranking of issues within the categories (treatment side effects, disease symptoms, quality of life) was produced based on this total.

### Assessing Measurement Tools

Measurement tools identified in the literature search were assessed using a published structure for assessing PCOM for use in clinical trials by Fitzpatrick et al.^15^ This was based on assessing properties of; reliability, validity, responsiveness, precision, interpretability, appropriateness, acceptability, and feasibility for each HR-QoL tool (full definitions of these terms are provided in the Supplement).

Reliability, validity and responsiveness were assessed using the results of a systematic review previously published by Khan et al on the use of quality of life measurement in interventional TB research for SF-36, WHOQOL BREF, SGRQ, MOS-HIV, and FACIT-TB.^12^ Evidence in other respiratory diseases for SF-12, SF-6, and EQ-5D-5L was obtained through a literature search (see Supplement).

Precision and interpretability of the measurement tool scores were judged by researchers with expertise in clinical trials, statistical methodology, and person-focussed research. The panel individually reviewed the definitions for the terms “precision” and “interpretability” and agreed gradings for the different measurement tools with “good” defined as the instrument satisfying the criteria for that field with no changes needed, “fair” indicating that the tool would require some changes to satisfy all the criteria for that field, and “poor” defined as the instrument failing to satisfy the criteria for that field even with some changes made. The measurement tools were then assessed by these researchers as a panel and gradings agreed.

Appropriateness and acceptability of the measurement tools for use in TB trials were assessed by trained TB Community Advisory Group (CAG) members from the UNITE4TB Consortium. The results of the ranked issues relating to treatment, disease symptoms and quality of life from the focus groups were presented to the CAG members along with the characteristics of each measurement tool at an online workshop. Each of these fields was rated as “good”, “fair” or “poor” using the previously agreed definitions.

The feasibility of administering the measurement tool to trial participants was assessed by site staff from South Africa and Mongolia. The site staff were asked to rate the measurement tools using the same “good”, “fair”, or “poor” criteria after a review of each tool’s characteristics with descriptors provided for each of these ratings.

## Results

### Literature Review

The literature search for HR-QoL measures used in TB research yielded eight candidate tools previously used in interventional studies as presented in **Table 1**, and only one of these was disease-specific for TB.^14^ Systematic reviews of health-related quality of life measures used as endpoints in TB studies^12^ and also more generally in disease assessment^10^ were also identified.

**Table 1.** Summary of HR-QoL scores evaluated.

| HR-QoL Assessment Tool | Domains | Timeframe | Scoring System |
| --- | --- | --- | --- |
| **SF-36** | Vitality (or energy)<br>Physical functioning<br>Bodily pain<br>General health perceptions<br>Physical role functioning<br>Emotional role functioning<br>Social role functioning<br>Mental health or emotional wellbeing | Last seven days | 8 scaled scores, which are weighted sums of their section<br>Each scale is directly put into a 0-100 scale |
| **WHOQOL BREF** | Physical health<br>Psychological<br>Social relationships<br>Environment | Last two weeks | Response levels vary across questions<br>Scores for individual domains only |
| **SGRQ** | Symptoms<br>Activity<br>Impacts | Current status | Response levels vary across questions<br>Scores for domains and total score |
| **MOS-HIV** | Health perceptions<br>Pain<br>Physical<br>Role functioning<br>Social functioning<br>Cognitive functioning<br>Mental health<br>Energy<br>Health distress<br>Quality of life | "Typical day" (for some questions) or last four weeks (for some questions) | Response levels vary across questions<br>Domain scores and summary scores for physical/mental health |
| **FACIT-TB** | Physical Well-being<br>Social/Family Well-being<br>Emotional Well-being<br>Functional Well-being<br>Spiritual Well-being | Last seven days | Ordinal categorical responses: Not at all/ A little bit/ Some-what/ Quite a bit/ Very much<br>Domain and Total scores |
| **SF-12** | Vitality (or energy)<br>Physical functioning<br>Bodily pain | Last seven days | Ordinal categorical responses with associated numerical score: Not at all/ A little bit/ Some-what/ Quite a bit/ Very much |
|  | General health perceptions<br>Physical role functioning<br>Emotional role functioning<br>Social role functioning<br>Mental health or emotional wellbeing |  | Scores for the physical and mental components |
| **SF-6** | Vitality (or energy)<br>Physical functioning<br>Bodily pain<br>General health perceptions<br>Physical role functioning<br>Emotional role functioning<br>Social role functioning<br>Mental health or emotional wellbeing | Last seven days | Ordinal categorical responses with associated numerical score: Not at all/ A little bit/ Some-what/ Quite a bit/ Very much<br>Overall total score calculated |
| **EQ-5D-5L** | Mobility<br>Self Care<br>Pain<br>Usual activities<br>Psychological status<br>Overall health state | Day of questionnaire | Ordinal categorical responses within domains that map to different levels (1 -> 5)<br>Health state calculated from levels across domain and overall state from visual scale |

Health-related quality of life (HR-QoL) assessment tools used in TB interventional studies identified from a literature search with domains covered, timescale assessed and scoring approach presented

### First Round of Focus Groups

The first round of focus groups included nine groups held across the three sites involving 57 participants. The demographics for the participants can be found in **Table 2**, and 93% had received bedaquiline-containing regimens as their treatment for TB. The qualitative work used to determine the main themes relating to treatment and disease has been published elsewhere.^11^

**Table 2.** Focus group participant characteristics.

|  |  | Mongolia | South Africa | Georgia |
| --- | --- | --- | --- | --- |
| Focus Group 1 | Male Sex (%n) | 5<br>(33%) | 11<br>(55%) | 15<br>(68%) |
|  | Mean Age (Range) | 39<br>(30 – 65) | 41<br>(26 – 61) | 37<br>(19 – 67) |
|  | Employed (%n) | 8<br>(53%) | 4<br>(20%) | 16<br>(73%) |
|  | HIV-positive (%n) | 0<br>(0%) | 15<br>(75%) | 0<br>(0%) |
|  | TB Treatment Completed (%n) | 3<br>(20%) | 14<br>(70%) | 22<br>(100%) |
| Focus Group 2 | Male Sex (%n) | 11<br>(69%) | 5<br>(50%) |  |
|  | Mean Age (Range) | 43<br>(23 - 61) | 37<br>(26 - 53) |  |
|  | Employed (%n) | 5<br>(31%) | 4<br>(40%) |  |
|  | HIV-positive (%n) | 0<br>(0%) | 4<br>(40%) |  |
|  | TB Treatment Completed (%n) | 2<br>(13%) | 7<br>(70%) |  |

Characteristics of participants in two rounds of focus groups in Mongolia, South Africa, and Georgia. The numbers presented are totals across all focus groups run at the site as part of that round. For logistical reasons, the Georgia site was unable to participate in the second round of focus groups

The group work generated a list of items relating to treatment and disease symptoms. These items are presented in **Table 3** and were grouped under the headings “overall quality of life”, “disease symptoms”, and “treatment side effects”.

**Table 3.**
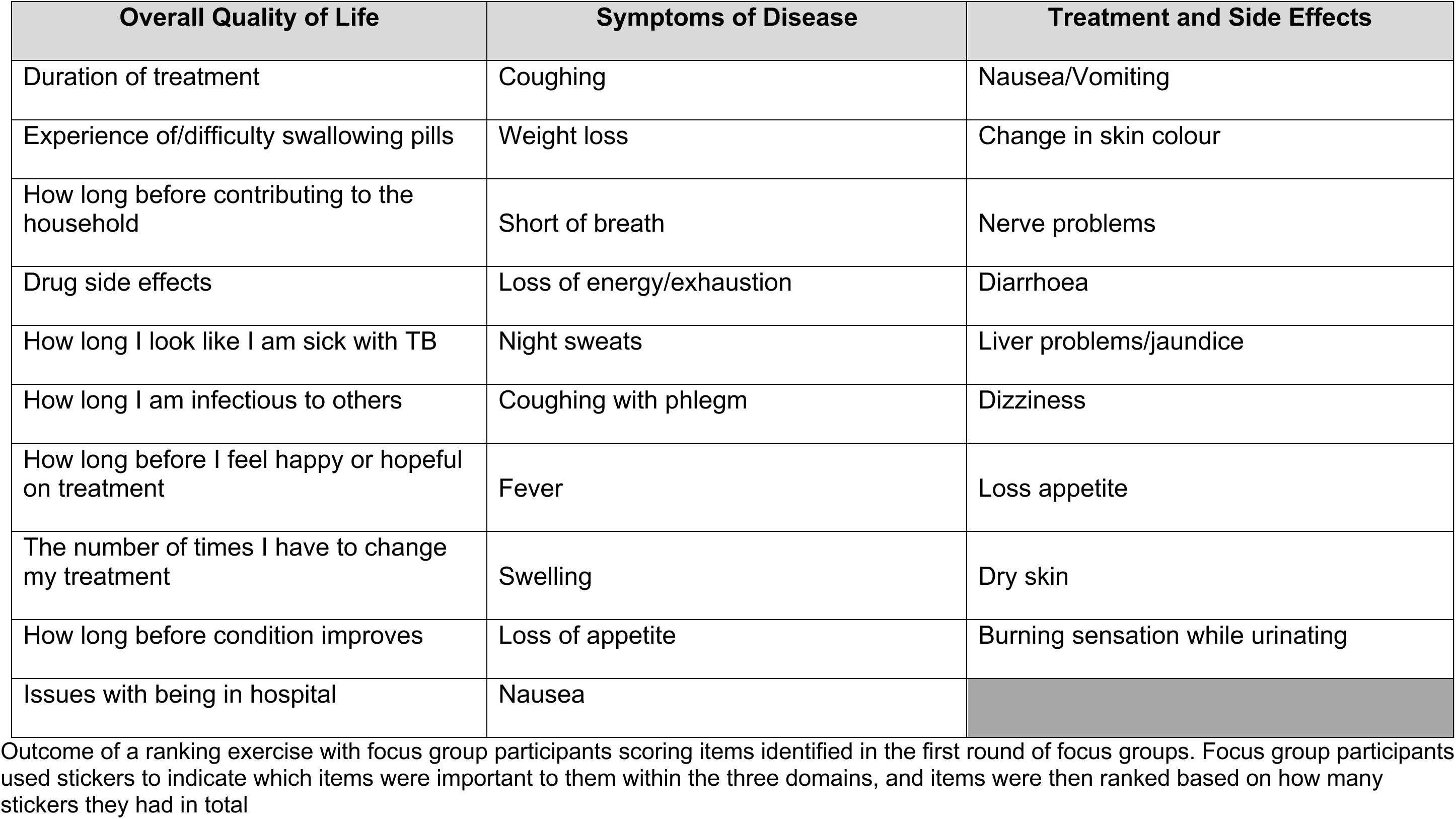
Items ranked in Round 2 of the focus groups by participants with highest-ranked at top.

### Second Round of Focus Groups

There were 26 participants involved in four focus groups at the sites in Mongolia and South Africa, and their demographics are presented in **Table 2**. The highest ranked issues relating to overall treatment experience and quality of life were duration of treatment, experience of swallowing pills, and how long before being able to contribute to the household. The symptoms reported as being most important were coughing, weight loss and breathlessness. Side effects related to treatment that were given the highest rankings were nausea/vomiting, change in skin colour, and nerve problems.

A full list of the issues ranked according to individual focus groups and totals across all sites can be found in the supplement.

### Assessing the Measurement Tools

**Table 4** summarises the results for the evaluation of reliability, validity, and responsiveness using the results of a systematic review(ref) and separate literature search. More detail on the evaluation can be found in **Section S2** in the Supplement, including breakdowns of site staff assessment of feasibility and references from the systematic review and literature search.

**Table 4.** Assessment of outcome measures using systematic review data and scoping review of literature.

|  | SF-36* | WHOQOL BREF* | SGRQ* | FACIT-TB* | MOS-HIV* |  | SF-12§ | SF-6§ | EQ-5D-5L§ |
| --- | --- | --- | --- | --- | --- | --- | --- | --- | --- |
| **Appropriateness\*\*** | Poor | Poor | Fair | Good | Poor |  | Poor | Poor | Poor |
| **Reliability** | Excellent | Good<br>Excellent internal consistency<br>Good test-retest reliability | Poor internal consistency<br>Fair reliability | Excellent | Fair |  | Fair | Fair | Good |
| **Validity** | No evidence on validity in TB | Good content validity<br>Fair structural validity | Good content validity<br>No evidence on structural validity | Good | Good content validity<br>Poor structural validity |  | Good | Good | Good |
| **Responsiveness** | No evidence on responsiveness in TB | No evidence on responsiveness in TB | No evidence on responsiveness in TB | Excellent | No evidence on responsiveness in TB |  | Fair | Good | Good |
| **Precision†** | Excellent | Excellent | Excellent | Excellent | Excellent |  | Fair | Poor | Good |
| **Interpretability of scores†** | Fair | Fair | Good | Fair | Good |  | Fair | Fair | Good |
| **Acceptability to participants\*\*** | Poor | Poor | Fair | Fair | Good |  | Poor | Poor | Good |
| **Feasibility‡**<br>**(judged by site facilitators)** | Poor | Poor | Fair | Fair | Fair |  | Fair | Fair | Fair |

\* Evidence on reliability, validity and responsiveness obtained from systematic review<sup>14</sup>; § Evidence on reliability, validity and responsiveness obtained from scoping review of literature; \*\* Judged by community panel; † Judged by authors; ‡ Judged by site staff (poorest rating from site staff shown)

Reliability was found to be excellent or good for SF-36, WHO BREF, FACIT-TB, and EQ-5D-5L. However, it was “poor” for the SGRQ, and “fair” for MOS-HIV, SF-12, and SF-6. In all of the measurement tools validity was rated as at least “good” or “fair” with the exception of MOS-HIV, which was “poor” for structural validity, and SF-36 for which no evidence was available in TB disease. Responsiveness was judged as “excellent” for FACIT-TB, “good” for SF-6 and EQ-5D-5L, and “fair” for the SF-12. There was no evidence in TB disease found for SF-36, WHO BREF, SGRQ and MOS-HIV in the systematic review.^12^ Precision was judged as “excellent” or “good” for all tools, except for SF-12 and SF-6 which were considered “fair” and “poor”, respectively.

Interpretability was judged as “good” for SGRQ, MOS-HIV, and EQ-5D-5L; all other measurement tools were rated as “fair” by the panel of researchers with experience in tuberculosis and clinical trials. For acceptability, the CAG panel judged EQ-5D-5L and MOS-HIV as being “good”, while SGRQ and FACIT-TB were considered “fair”. Acceptability was considered “poor” for SF-36, SF-12, SF-6, and WHOQOL BREF. In relation to the priorities set by the focus groups the CAG panel judged SF-36, SF-12, SF-6, EQ-5Q-5L, WHOQOL BREF, and MOS-HIV as “poor” in respect to appropriateness for use in a DR-TB clinical trial. FACIT-TB and SGRQ were judged to be “good” and “fair” respectively in terms of appropriateness .

The site staff in South Africa and Mongolia considered MOS-HIV to be “fair” in terms of its feasibility, but there was a lack of agreement for the other measurement tools; the lowest ratings are presented in **Table 4**. All the tools were considered to be either “good” or “fair” by the sites, with the exception of SF-36 and WHOQOL BREF as both were considered “poor” by South Africa because of difficulty understanding and time to complete, respectively. FACIT-TB was rated as “fair” by South Africa, because it was thought to be not easy to understand, and “good” by Mongolia’s site staff as they felt the questionnaire was straightforward and short.

FACIT-TB was the only measurement tool assessed that was rated “excellent”, “good” or “fair” in all of the fields following assessment by community representatives, researchers and clinical trial site staff specialising in TB and use of results from a systematic review.

## Discussion

This work sought to identify one or more measurement tools that could be used to collect data on disease and treatment-specific participant priorities for use in clinical trials of treatments for tuberculosis. A literature search identified candidate tools, focus group work elicited ranked priorities, and the most appropriate measurement tool was then selected using a published assessment framework.^15^ Following on from this assessment involving TB survivors, community advocates, researchers, and academic TB physicians this work has identified FACIT-TB is the most useful tool available to capture priorities relating to both disease and treatment experience during ongoing TB treatments.

While the work was conducted focussing on people with lived experience of DR-TB, the FACIT-TB score would be appropriate for use in both DS-TB and DR-TB as the domains assessed relate to experience of disease and side effects from treatment. This is particularly relevant as we move into an era where people with both DS-TB and DR-TB are being recruited into the same trial where completely novel regimens are being investigated. In many cases, these regimens do not contain the drugs that are the basis for traditional labels of drug-susceptibility (e.g. rifampicin), and these definitions could be out-dated.

We have shown that among identified priorities relating to DR-TB and its treatment, some are considered more important than others. Duration of treatment, experience with pills, and how long it is until being able to contribute to the household were the most important overall quality of life measures. Cough, weight loss and breathlessness were the most important symptoms of disease, and nausea/vomiting, changes in skin colour, and nerve problems were the highest ranked side effects. Previous work has demonstrated an association between priorities of people with TB relating to treatment and their adherence to treatment, including addressing side effects, pill burden and functional status.^17–21^ A failure to capture these priorities at an early stage of investigating novel treatments would mean running a risk of masking barriers to their real-world use, and approving regimens that would ultimately represent small gains due to poor tolerance. The current goal in TB therapeutics research is to identify regimens that are effective, simple and well-tolerated and capturing these priorities is intrinsically a part of this.

Outcome measures in clinical trials require careful thought, and are one of the components that make up the trial’s estimand. A failure to choose the correct outcome can mean that the trial’s research question is not answered, regardless of the other elements of the trial’s design.^21^ This evaluation of the measurement tools by community representatives as part of this project has shown that these tools are not interchangeable, and some were even thought to be inappropriate for use in a TB trial. The FACIT-TB tool^22^ captures aspects of disease and treatment relating to physical, emotional, and social priorities for people being treated for TB in a clinical trial. It was judged appropriate for use by an experienced CAG based on priorities elicited from focus groups of people with lived experience, and by capturing subjective experience numerically it allows for quantitative analysis to be performed of experience relating to disease and treatment. A systematic review of its use in interventional trials confirmed that it would meet the criteria set out by the COSMIN Initiative for reliability, validity and responsiveness.^14^ This measurement tool would help ameliorate many of the existing issues, if it were to be used as the standard PCO for DR-TB clinical trials, by facilitating comparison between different novel treatment regimens in both trials and meta-analysis.

The evaluation of the PCOs was robust in this project. The selection of FACIT-TB^22^ as the most appropriate disease-specific PCO was based on evaluations by TB survivors and clinical research staff with experience of TB clinical trials. The evaluation was undertaken using a recognised approach for assessing tools for clinical research.^15^ However, there are still considerations relating to the use of outcome measures that would need to be considered prior to it being considered as a standard approach for evaluating the participant experience of novel treatment regimens. First is the need for a PCO to be validated in multiple languages, and in the case of FACIT-TB it is currently only validated for use in a limited number of languages, although work is ongoing to improve this (https://www.unite4tb.org/). Second, the use of the score will need evaluation to determine how best to obtain meaningful information relating to the participants’ experience. Evaluation of quality of life over the course of treatment will involve consideration of issues relating to the handling of longitudinal data.^23^ ^24^ In particular, how frequently should the questionnaire be applied and how should repeated measurements be interpreted (e.g. change from baseline, rate of change, or final result only)? Also, should there be a cut-off value for the lowest permitted score (or drop in score) allowed while on treatment for the regimen to be considered “acceptable”?

Third, the use of PCOMs also requires consideration around how they are evaluated in light of the other characteristics of the regimen under investigation. The issue of how to utilise PCOM results in the context of efficacy and safety results is under review by the FDA at the time of writing,^25^ and would need careful thought with input from TB survivor representatives. For example, what is the correct method for interpreting trial results that demonstrate cure rates of 95% after four weeks of therapy with an acceptable safety profile but poor scores associated with the chosen PCOMs? While this work has identified the most suitable tool for capturing participant priorities in clinical trials, this tool must be used correctly to ensure data are collected in a meaningful way.

This study has limitations. Firstly, this work was carried out with people who have experience of DR-TB, and there should be some caution around generalising to TB disease and its treatment more generally in particular those with asymptomatic TB who may have different concerns. While multiple sites in three continents were used, the work cannot claim to represent perspectives beyond the regions included. The final evaluation of the person-assessed “appropriateness” of the tools, which led to the exclusion of any that were judged inappropriate, was carried out by a small number of community representatives. Further, FACIT-TB may not cover all items identified as important for specific trial populations.

Notably aspects such as medication acceptance and tolerability could be captured using different tools. Trial teams should determine with the input of community advisory boards additional PCOM relevant to their trials. In addition to disease specific PCOM such as FACIT-TB there may be other reasons to include more generic tools such as EQ-5D and SF-36 which may be required for health economic analysis.

In conclusion, this work has found that the FACIT-TB score was the most appropriate disease specific measurement tool to be used as a PCOM in a TB clinical trial. FACIT-TB will capture priorities relating to both TB disease and treatment, but how best to use the score in terms of repeated measures and their interpretation needs to be determined.

Further work is also needed to define where the score should lie in the analysis plan, e.g. as a co-primary endpoint with efficacy or secondary endpoint, how will poor PCOM scores be interpreted in the context of high efficacy? However the score is ultimately used in trials, adopting a standard approach to assessing priorities of people with TB will facilitate data synthesis and meta-analysis as well as producing PCO results that are more clear and interpretable for individual trials. Addressing these priorities are an essential part of developing any new intervention and there is a need for PCOs in TB clinical trials to be given more prominence if the regimens being investigated are to be successfully integrated into real-world practice.

## Data Availability

All data produced in the present study are available upon reasonable request to the authors

## Acknowledgements

We would like to acknowledge Dr Parveen Dhesi for her work in qualitatively analysing the outputs from the first round of focus groups.

## References

1. Global tuberculosis report 2024. Geneva: World Health Organization; 2024.

2. Reid M, Agbassi YJP, Arinaminpathy N, Bercasio A, Bhargava A, Bhargava M et al. Scientific advances and the end of tuberculosis: a report from the Lancet Commission on Tuberculosis. Lancet. 2023;402:1473–98

3. Conradie F, Bagdasaryan TR, Borisov S, Howell P, Mikiashvili L, Ngubane N et al. Bedaquiline-Pretomanid-Linezolid Regimens for Drug-Resistant Tuberculosis. N Eng J Med. 2022;387:810–823

4. Goodall RL, Meredith SK, Nunn AJ, Bayissa A, Bhatnagar AK, Bronson G et al. Evaluation of two short standardised regimens for the treatment of rifampicin-resistant tuberculosis (STREAM Stage 2): an open-label, multicentre, randomised, non-inferiority trial. Lancet. 400;1858–68

5. Nyang’wa BT, Berry C, Kazounis E, Motta I, Parpieva N, Tigay Z et al. A 24-Week, All-Oral Regimen for Rifampin-Resistant Tuberculosis. N Eng J Med. 2022;387:2331 – 43

6. Cevik M, Thompson LC, Upton C, Rolla VC, Malahleha M, Mmbanga B et al. Bedaquiline-pretomanid-moxifloxacin-pyrazinamide for drug-sensitive and drug-resistant pulmonary tuberculosis treatment: a phase 2c, open-label, multicentre, partially randomised controlled trial. Lancet Infect Dis. 2024;24:1003–1014

7. Alonso-Coello P, Oxman AD, Moberg J, Brignardello-Petersen R, Akl E, et al. GRADE Working Group. GRADE Evidence to Decision (EtD) frameworks: 2. Clinical practice guidelines. BMJ. 2016;353:i2089

8. Guidance on evidence generation on new regimens for tuberculosis treatment. Geneva: World Health Organization; 2024.

9. Kruijshaar ME, Lipman M, Essink-Bot ML, Lozewicz S, Creer D, et al. Health status of UK patients with active tuberculosis. Int J Tuberc Lung Dis. 2010;14: 296–302

10. Atif M, Sulaiman SA, Shafie AA, Asif M, Sarfraz MK, et al. Impact of tuberculosis treatment on health-related quality of life of pulmonary tuberculosis patients: A follow-up study. Health Qual Life Outcomes. 2014;12: 19

11. Mamani M, Majzoobi MM, Ghahfarokhi SM, Esna-Ashari F, Keramat F. Assessment of health-related quality of life among patients with tuberculosis in Hamadan, Western Iran. Oman Med J. 2014;29: 102–105

12. Brown J, Capocci S, Smith C, Morris S, Abubakar I, Lipman M. Health status and quality of life in tuberculosis. Int J Infect Dis. 2015;32:68–75

13. South A, Dhesi P, Tweed CD, Tsogt B, Staples S, Tukvadze N et al. Patient’s priorities around drug-resistant tuberculosis treatment: A multi-national qualitative study from Mongolia, South Africa and Georgia. Glob Public Health. 2023;18:2234450

14. Khan S, Tangiisuran B, Imtiaz A, Zainal H. Health Status and Quality of Life in Tuberculosis: Systematic Review of Study Design, Instruments, Measuring Properties and Outcomes. Health Sci J. 2017;11:484

15. Fitzpatrick R, Davey C, Buxton MJ, Jones DR. Evaluating patient-based outcome measures for use in clinical trials. Health Tech Ass. 1998;2(14)

16. Abdulelah J, Sulaiman SAS, Hassali MA, Blebil AQ, Awaisu A, Bredle JM. Development and Psychometric Properties of a Tuberculosis-Specific Multidimensional Health-Related Quality-of-Life Measure for Patients with Pulmonary Tuberculosis. Val Health Reg Iss. 2015;6:53–59

17. Kaona FAD, Tuba M, Siziya S, Sikaona L. An assessment of factors contributing to treatment adherence and knowledge of TB transmission among patients on TB treatment. BMC Pub Health. 2004;4:68

18. Tola HH, Tol A, Shojaeizadeh D, Garmaroudi G. Tuberculosis Treatment Non-adherence and Lost to Follow-up Among TB Patients with or Without HIV in Developing Countries: A Systematic Review. Iran J Pub Health. 2015;44:1–11

19. Gebremariam MK, Bjune GA, Frich JC. Barriers and facilitators of adherence to TB treatment in patients on concomitant TB and HIV treatment: a qualitative study. BMC Pub Health. 2010;10:651

20. Woimo TT, Yimer WK, Bati T, Gesesew HA. The prevalence and factors associated for anti-tuberculosis treatment non-adherence among pulmonary tuberculosis patients in public health care facilities in South Ethiopia: a cross-sectional study. BMC Pub Health. 2017;17:269

21. European Medicines Agency. ICH E9 (R1) addendum on estimands and sensitivity analysis in clinical trials to the guideline on statistical principles for clinical trials. 2020

22. Dujaili J.A., Syed Sulaiman S.A., Hassali M.A., Blebil A.Q., Awaisu A., Bredle J.M. Development and psychometric properties of a tuberculosis-specific multidimensional health-related quality-of-life measure for patients with pulmonary tuberculosis. Value in Health Regional Issues 6C (2014): 53–59

23. Bascoul-Mollevi C, Barbieri A, Bourgier C, Conroy T, Chauffert B, Hebbar M et al. Longitudinal analysis of health-related quality of life in cancer clinical trials: methods and interpretation of results. Qual Life Res. 2020;30:91–103

24. Donneau AF, Mauer M, Coens C, Bottomley A, Albert A. Longitudinal quality of life data: a comparison of continuous and ordinal approaches. Qual Life Res. 2014;23:2873–2881

25. FDA. Patient-focused Drug Development: Incorporating Clinical Outcome Assessments Into Endpoints for Regulatory Decision-making (DRAFT). 2023

