## Supplemental Material for "Evaluation of person-centred outcome measures for use in clinical trials of tuberculosis therapeutics"

### Evaluation of person-centred outcome measures (PCOMs) for use in clinical trials of tuberculosis therapeutics – Supplementary Material

#### S1 – Definitions used in assessing PCOMs

##### Appropriateness: Is the content of the instrument appropriate to the questions which the clinical trial is intended to address?

###### Summary

In more general terms, appropriateness of an instrument for a trial will involve considering the other criteria we have identified and discuss below; evidence of reliability, feasibility, and so on. In the more specific terms with which we have summarised the rather disparate literature on appropriateness, the term requires that investigators consider as directly as possible how well the content of an instrument matches the intended purpose of their specific trial.

##### Reliability: Does the instrument produce results that are reproducible and internally consistent?

###### Summary

Overall, we are here concerned with how reproducible an instrument is, and, where relevant, how internally consistent items are in scales. Reproducibility can be assessed by fairly specific methods so that it is a relatively straightforward aspect of an instrument to assess when such evidence is available. In reality, internal consistency is more frequently reported although very high internal consistency is not always considered desirable.

#### Validity: Does the instrument measure what it claims to measure?

##### Summary

The apparently simple question as to whether an instrument measures what it purports to measure has to be considered by means of a range of different kinds of evidence including how content was determined, inspection of the content, and of patterns of relationships to other variables. Because no single set of observations is likely to determine validity and different kinds of evidence are needed, judgement of this property of an instrument in relation to a specific trial is not straightforward.

#### Responsiveness: Does the instrument detect changes over time that matter to patients?

##### Summary

The need for an instrument to be responsive to changes that are of importance to patients should be of evident importance in the context of clinical trials. Whilst there are no universally agreed methods for assessing this property, at a more general level all discussions require evidence of statistically significant change of some form from observations made at separate times and when there is good reason to think that changes have occurred that are of importance to patients.

#### Precision: How precise are the scores of the instrument?

##### Summary

Overall, we are here concerned with how precise are the distinctions made by an instrument, with at one extreme instruments that make very few rather gross distinctions between levels of health and illness and, at the other extreme, instruments that make many more specific distinctions. Given that clinical trials are frequently concerned with looking for difficult-to-detect differences between treatments, it might appear that the capacity to make numerous distinctions is in itself desirable. However, the literature has suggested a number of ways in which this would be misguided and not reflect accurate precision.

#### Interpretability: How interpretable are the scores of an instrument?

##### Summary

Interpretability is concerned with how meaningful are the scores from an instrument To date, it is not possible to compare patient-based outcome measures in terms of how interpretable developers have managed to make their instruments, although clearly those instruments that are more regularly included in trials and population studies will come to be more widely known and more familiar by use (Greenfield and Nelson, 1992).

#### Acceptability: Is the instrument acceptable to patients?

##### Summary

Evidence is required that an instrument is acceptable to patients. The simplest and most direct form of such evidence is that it has consistently been associated with high response rates. Early on in the development of an instrument, this property may have been more directly tested by eliciting views of patients about the instrument.

### Feasibility: Is the instrument easy to administer and process?

#### Summary

The time and resources required to collect, process and analyse a patient-based outcome measure are not often independently reported so that evidence may not be readily available. A judgement of this aspect of an instrument has to be made in the context of clinical trials given that this will be but one component of burden on participants that will determine the overall viability of a trial and therefore the quality of its final results.

### S2 – Site staff assessments of feasibility for PCOMs

| Outcome measure | Feasibility of administering outcome measure: Mongolia | Key reason for your answer: Mongolia | Feasibility of administering outcome measure: S Africa | Key reason for your answer: S Africa |
|---|---|---|---|---|
| **EQ5D** | Good | This questionnaire is easy to administer and takes a short time to complete, because it's simple for the patients to understand and complete themselves. | Fair | Section on depression and anxiety is hard and needs extra explanation. Not sure that 0-100 scale works well, Suzanne has experience of patients scoring as 99 throughout even when very ill! (This could be a population-specific issue because clinic days may be better than average days for the patients attending.) |
| **SF-36** | Poor | It would be difficult for patients to understand; the scored answers seem complicated to understand or distinguish, for example what is "a good bit of time", "some of time" etc. True or false statements are also not good, as most of patients will be truly ill. | Fair | Long questionnaire. Questions requiring recall over a 4-week period can be hard to answer if the patient's experience of e.g. pain has varied over that period, so the person delivering the questionnaire needs specialist training. |

| | | | | |
|---|---|---|---|---|
| **SF-12** | Fair | This is easier to complete, but some of the examples in the question (2a) would be better to change; pushing table and playing bowling are the same moderate activities | Good | Same issue as SF36 over questions requiring recall over a 4-week period, but the questionnaire is short. |
| **SF-6** | Fair | Options are not easy to understand or even to translate. | Good | Same issue as SF36 over questions requiring recall over a 4-week period, but the questionnaire is short. |
| **WHOQOL BREF** | Poor | It would take quite time to complete for the patients, as well as for the clinic staff to explain | Fair | Some wording of questions is unnatural and needs explaining e.g. “What is your quality of life?”. Some sensitive questions about finance and living conditions. These would result in more time spent with trial participant, increased workload for site etc |
| **SGRQ** | Fair | Easier to complete, but if instead of True or False to amend into Yes, No would be better | Good | Straightforward to implement. Lots of questions with binary True/False answers, not too many options for patients to think |

| | | | | |
|---|---|---|---|---|
| | | | | about. Not very long. Questions don't seem to cover sensitive topics. |
| **FACIT-TB** | Fair | Not easy to complete, the layout of questions and options are not easy to understand, thus will require someone to explain | Good | Questions look straightforward and short. Length of questionnaire is ok because the questions are closely linked to TB and patients' experiences. |
| **MOS-HIV instrument** | Fair | Would be good to have fewer multiple answer scored questions | Fair | Questionnaire feels long. Some terminology is unnatural and needs extra explanation e.g. "full of pep", "downhearted and blue". Language isn't always culturally appropriate. |

### S3 – Facilitator guide for first round of focus groups

**Background to the study:**
Drug-resistant tuberculosis (DR-TB) is a global health emergency and is one of the major drivers behind the on-going tuberculosis pandemic. Several new treatment regimens are currently under evaluation in different clinical trials, but clinicians working in the field will need to know how to compare these treatments against each other when managing individual patients. As part of this, clinicians will need to know what is important to patients about their treatment. This project will use focus groups and structured interviews with people who have been treated for DR-TB at sites in South Africa, Mongolia, Georgia, and Brazil to explore patient views around treatment. Over the course of the work we will:

1. Identify the aspects of treatment most important to DR-TB patients through convening focus groups and application of qualitative research methods
2. Develop specific clinical trial outcome measures that reflects patient-identified outcome of interest by modifying existing tools and simulation.
3. Develop wider consensus through convening a wider group of stakeholders to agree on the patient-centred outcome measure

At the end of this project we will have developed outcome measures that reflect what is important to patients for use in a novel trial design to rank new DR-TB regimens. Such a trial will inform national treatment policy, clinician treatment choices, and facilitate patient involvement in informed decision-making. Ultimately this will lead to better outcomes and help global efforts to end the TB epidemic.

**Who is being recruited:**
We aim to recruit 20-30 patients from each site. Focus group discussions and interviews will involve participants who are 18 years or older, culture-negative for tuberculosis, and who are well enough to participate. They should have either completed treatment for DR-TB in the last 12 months, or be in the "continuation phase" of their treatment (or equivalent). Patients with signs or symptoms of active disease will not be able to participate.
Participants will be recruited by the site staff actively approaching them and efforts will be made to ensure that focus groups are representative of the range of DR-TB patients in terms of ethnicity, economic standing, and treatment experience.

**Who will be doing the research:**
This project is being conducted by researchers from the MRC Clinical Trials Unit at UCL, London, UK in partnership with the following:

- UCL Institute for Global Health
- National Centre for Communicable Diseases, Ulaanbaatar, Mongolia
- TB and HIV Investigative Network (THINK), Durban, South Africa
- National Center for Tuberculosis and Lung Disease, Tbilisi, Georgia
- National Institute of Infectious Diseases Evandro Chagas, Fiocruz, Brazil. This site will only be involved in the second round of focus groups due to current pressures from COVID-19

**What this study will involve:**
The study will run between March and May 2021. Participants will take part in focus group discussions composed of approximately 8 individuals with 2 facilitators present. There will be two focus group sessions and structured interviews conducted with volunteers.

1. The **first focus group session** will use a combination of "story stems" relating to a particular aspect of treatment that the group will propose endings for and discuss further, as well as time for more general discussion around any topics that participants feel need to be raised.
2. A **second focus group** will be used to evaluate proposed clinical trial outcome measures based (1) & (2). Participants will assess the outcome measures in terms of what information they think would be gathered and whether this aligns with what they feel are patient priorities

**What information will be kept about participants:**
Participant information will be collected and stored by the local sites to allow them to recruit the participant and maintain contact for the duration of the study. Personal information such as name, date of birth, address, DR-TB treatment status, HIV status, and other defining characteristics will be securely stored by the site and destroyed at the end of the study.
Focus group work and structured interviews will be recorded and notes will be taken by the facilitators. The recordings will be transcribed and translated before being shared with the research team along with facilitator notes. Any identifying information will be removed prior to sharing the materials and secure online sharing platforms will be used.

**How participants dignity and safety will be protected:**
Participants can use their real name or a pseudonym, depending on their preference, during the group discussions. Complete anonymisation cannot be guaranteed, but participant names will not be recorded in the transcripts, focus group notes, or interview notes and instead will be replaced with an identifier. Informed consent will be obtained prior to participants taking part in focus groups or interviews.
Measures in keeping with local guidance will be applied to ensure risk from both tuberculosis and COVID-19 will be minimised for participants. This will include appropriate personal protective equipment (PPE), only involving participants who are culture negative, and symptom screening.
In the unlikely event that a participant tells the researchers information that they deem to represent a danger to the individual or someone else, the researchers may be compelled to contact a relevant medical professional.

**Focus Group 1: Story Stem Discussion**

Housekeeping activities at beginning of discussion:
- Ensure all participants feel ready to engage in the discussion (e.g. adequate PPE if in person and no technical issues if conducted via online platform)
- Highlight any relevant points for the venue (e.g. fire exits)
- Ensure that pens/pencils and paper are available for the participants
- Recording equipment should be tested prior to group
- Read through ground rules and introduction tract

**Ground rules:**
Thank you to everyone for agreeing to take part in this study and for volunteering your time. The time spent in the focus group discussion may involve talking about topics that are personal and potentially upsetting, and as such we have ground rules to ensure that we remain respectful throughout.
- There are no right answers, and what we are interested in exploring is personal opinions around the topics introduced
- We should remain respectful of each other during the discussion and if any participants consistently demonstrate behaviour that is unacceptable (for example, name-calling, an aggressive or dismissive attitude, or an argumentative stance) then they could be asked to leave by the facilitators

- No participants will be forced to contribute to any part of the discussion and are free to leave at any time. The intention to speak can be indicated by a raised hand (or similar) and the facilitator will work to provide space to speak in the discussion
- Everything that is said in the focus group is to remain confidential and personal opinions expressed in the discussion are not to be shared outside of the focus group be others

**Focus Group 1 Introduction:**
We will explore issues around drug-resistant tuberculosis in the focus group and the aim is to gather information about what patient priorities actually are in relation to treatment. This information will then be used to develop measures of treatment success to be used in clinical trials investigating new drug-resistant tuberculosis treatments.
You are invited to complete a story about a fictional character—this means that you read the opening sentences of a story and then discuss what happens next. There is no right or wrong way to complete the story, and you can be as creative as you like. We are interested in the many different stories that people can come up with in the groups. Don't spend too long thinking about what might happen next, just complete the story in whatever way seems to make sense to you. The group will use the story to talk around certain issues relating to treatment, and the facilitators may ask questions to direct the conversation if necessary.

**Story Stems:**

- Provide each participant with pencil/pen and paper, if present in person
- Begin recording after alerting participants
- Spend approximately 15 mins on each story stem
- One facilitator should take notes during the discussion to record topics of particular interest and to supplement the recording

Story Stem 1: Beginning of treatment (15 - 20mins)

[Patient] has been told they have drug-resistant tuberculosis after being unwell for several months. The doctor meets with them and their partner to explain that treatment will last for at least nine to 12 months, and that treatment will likely have side effects. The doctor explains the disease is curable but the treatment will need to be taken daily to achieve this and reduce infectiousness. After the appointment [patient] is quiet and their partner asks what is on their mind.

Initial question

- *Can we come up with a few sentences to say what happens next? What do you think is going through the patients mind*

Follow-up probes (guided by facilitator)

- *What is the impact on the patient's life due to DR-TB pre-treatment (symptoms, quality of life, work)? Which of these do they want/hope treatment to address the most? What important activities might they not be able to due to DR-TB*
- *How do you think they feel about the duration of treatment? What about pill burden*
- *What would [patient] be most worried about at the beginning of treatment? Which side effects would they be most worried about?*

- *How do we think [patient]'s family feel about his diagnosis and treatment? What will they be most worried about?*
- *What might be contributing to stigma of having DR-TB at this stage?*

**Story Stem 2: During treatment (15 - 20mins)**

After completing 4 months of treatment, [patient] attends for a clinic review and is told that the treatment has been working because there are no tuberculosis bacteria in their sputum and their weight is improving. The doctors say that they need to perform some tests to make sure there are no dangerous side effects from the treatment. There will be least 5-8 more months of treatment. [Patient] has been trying to make sure they take the medication every day but this hasn't been easy. After the appointment their partner turns to [patient] and comments that maybe life can go back towards normal now and [patient] could get back to work.

Initial question

- *Can we come up with a few sentences to say what happens next? what is going through [patient]'s mind?*

Follow-up probes (guided by facilitator)

- *How are they feeling about the symptoms of tuberculosis disease at this point? Are they feeling better like life back to normal? What aspects of life might not be back to normal*
- *What might not have been easy about taking the medication? Do we think that [patient] is likely to keep taking their medication?*
- *What are some of the side effects they could be experiencing right now?*
- *How do they feel about the possibility of a treatment change? What if the duration of treatment were to change as well?*
- *What are the [patient]'s thoughts about the possible danger from the medication? How are monitoring visits affecting their life at this point in treatment?*

**Story Stem 3: End of treatment (15mins)**

[Patient] is nearing the end of treatment. At their last visit, the doctor told them the treatment has worked and their TB should be cured. They will need to attend for a final clinic review when they will have a chest X-ray done and be assessed. [Patient] is talking with their partner about the treatment and their plans for the future.

Initial question

- *Can we come up with a few sentences to say what happens next? What is going through [patient]'s mind and what will they be talking to their partner about?*

Follow-up probes (guided by facilitator)

- *Do you think they will have any concerns or on going issues (symptoms or side effects) after finishing treatment? Do you think the patient will feel like they are cured?*
- *How does their life now compare to their life before they became unwell with TB? Are there things that you think they will not do anymore that they were able to do before?*
- *How is their partner likely to react to the news?*
- *What are their thoughts about the DR-TB returning?*

### S4 – Facilitator guide for second round of focus groups

**Background to the study:**
Drug-resistant tuberculosis (DR-TB) is a global health emergency and is one of the major drivers behind the on-going tuberculosis pandemic. Several new treatment regimens are currently under evaluation in different clinical trials, but clinicians working in the field will need to know how to compare these treatments against each other when managing individual patients. As part of this, clinicians will need to know what is important to patients about their treatment. This project will use focus groups and structured interviews with people who have been treated for DR-TB at sites in South Africa, Mongolia, and Georgia to explore patient views around treatment. Over the course of the work we will:

1. Identify the aspects of treatment most important to DR-TB patients through convening focus groups and application of qualitative research methods
2. Develop specific clinical trial outcome measures that reflects patient-identified outcome of interest by modifying existing tools and simulation.
3. Develop wider consensus through convening a wider group of stakeholders to agree on the patient-centred outcome measure

At the end of this project we will have developed outcome measures that reflect what is important to patients for use in a novel trial design to rank new DR-TB regimens. Such a trial will inform national treatment policy, clinician treatment choices, and facilitate patient involvement in informed decision-making. Ultimately this will lead to better outcomes and help global efforts to end the TB epidemic.

**Who is being recruited:**
Focus group discussions will involve participants who are 18 years or older, culture-negative for tuberculosis, and who are well enough to participate. They should have either completed treatment for DR-TB in the last 12 months, or be in the "continuation phase" of their treatment (or equivalent). Patients with signs or symptoms of active disease will not be able to participate. For the second focus group participants will ideally not have participated in the first focus group but where this is a challenge to recruit participants involved in the first focus group may be involved and in this scenario maybe outside this 12-month window.
Participants will be recruited by the site staff actively approaching them and efforts will be made to ensure that focus groups are representative of the range of DR-TB patients in terms of ethnicity, economic standing, and treatment experience.

**Who will be doing the research:**
This project is being conducted by researchers from the MRC Clinical Trials Unit at UCL, London, UK in partnership with the following:

- UCL Institute for Global Health
- National Centre for Communicable Diseases, Ulaanbaatar, Mongolia
- TB and HIV Investigative Network (THINK), Durban, South Africa
- National Center for Tuberculosis and Lung Disease, Tbilisi, Georgia

**What this study will involve:**
Participants will take part in focus group discussions composed of approximately 8 individuals with 2 facilitators present. There will be two focus group sessions conducted with volunteers.

3. The **first focus group session** used a combination of "story stems" relating to a particular aspect of treatment that the group proposed endings for and discussed further, as well as time for more general discussion around any topics that participants felt needed to be raised.

4. The **second focus group** will be used to define the disease symptoms that participants believe are important. The second part of these focus groups will be spent prioritising items related to both TB disease symptoms and treatment to inform the development of patient-centred outcomes.

**What information will be kept about participants:**
Participant information will be collected and stored by the local sites to allow them to recruit the participant and maintain contact for the duration of the study. Personal information such as name, date of birth, address, DR-TB treatment status, HIV status, and other defining characteristics will be securely stored by the site and destroyed at the end of the study.
Focus group work will be recorded and notes will be taken by the facilitators. The recordings will be transcribed and translated before being shared with the research team along with facilitator notes. Any identifying information will be removed prior to sharing the materials and secure online sharing platforms will be used.

**How participants' dignity and safety will be protected:**
Participants can use their real name or a pseudonym, depending on their preference, during the group discussions. Complete anonymisation cannot be guaranteed, but participant names will not be recorded in the transcripts, focus group notes, or interview notes and instead will be replaced with an identifier. Informed consent will be obtained prior to participants taking part in focus groups or interviews.
Measures in keeping with local guidance will be applied to ensure risk from both tuberculosis and COVID-19 will be minimised for participants. This will include appropriate personal protective equipment (PPE), only involving participants who are culture negative, and symptom screening.
In the unlikely event that a participant tells the researchers information that they deem to represent a danger to the individual or someone else, the researchers may be compelled to contact a relevant medical professional.

**Focus Group 2: Confirming important items and ranking by patients**

Housekeeping activities at beginning of discussion:

- Ensure all participants feel ready to engage in the discussion (e.g. adequate PPE if in person and no technical issues if conducted via online platform)
- Highlight any relevant points for the venue (e.g. fire exits)
- Ensure that pens/pencils and paper are available for the participants
- Recording equipment should be tested prior to group
- Read through ground rules and introduction tract

**Ground rules:**
Thank you to everyone for agreeing to take part in this study and for volunteering your time. The time spent in the focus group discussion may involve talking about topics that are personal and potentially upsetting, and as such we have ground rules to ensure that we remain respectful throughout.

- There are no right answers, and what we are interested in exploring is personal opinions around the topics introduced
- We should remain respectful of each other during the discussion and if any participants consistently demonstrate behaviour that is unacceptable (for example, name-calling, an aggressive or dismissive attitude, or an argumentative stance) then they could be asked to leave by the facilitators

- No participants will be forced to contribute to any part of the discussion and are free to leave at any time. The intention to speak can be indicated by a raised hand (or similar) and the facilitator will work to provide space to speak in the discussion
- Everything that is said in the focus group is to remain confidential and personal opinions expressed in the discussion are not to be shared outside of the focus group be others

**Focus Group 2 Introduction:**
We have recently conducted focus group discussions with DR-TB patients from South Africa, Mongolia and Georgia. These patients told us about their experience of TB treatment and disease. The aims of today's focus group are to make sure we have identified all the important aspects of treatment and the experience of TB disease after these first focus groups, and then try to arrange these points in order of importance for TB patients.

In addition the last focus group discussion had an emphasis on participants' experiences of treatment. Their initial experiences of disease were less fully explored. Understanding which symptoms of disease improve with treatment is important for this project, so we wish to further explore this in this second focus groups.

**Activity 1: Exploring initial experience of disease and the initial treatment response (15mins)**

- Provide each participant with pencil/pen and paper, if present in person
- Begin recording after alerting participants
- One facilitator should take notes during the discussion to record topics of particular interest and to supplement the recording

Initial questions/probes

- *Thinking back to when you first developed symptoms of TB. Which were the symptoms that troubled you most or that you found most bothersome?*
- *Which symptoms affected your quality of life the most?*
- *What symptoms affected your ability to work?*
- *What symptoms affected your ability to socialise?*
- *What symptoms caused you to seek healthcare?*
- *Once starting treatment what improvements did you first notice? How quickly did this improvement occur? How did you feel about this improvement?*
- *Overall what was worse for you the symptoms of TB or the side effects of the medication. Were you able to distinguish between them?*

**Activity 2: Confirming important aspects of treatment and disease (40mins)**

- Print out the 9 cards provided with items relating to TB disease and treatment prior to the focus group. These cards will have an associated picture to assist participants with limited reading ability
- Each participant in the group is provided with a set of cards (9 cards with text and picture, and 3 blank cards) and 15 small stickers

*Introduction to Activity 2 for participants:*
We will begin by showing everyone a list of different points relating to drug-resistant TB treatment that seemed to be important to the participants in the last focus group, and the aim of this activity is to try to decide how important each one is based on the group's opinion. These include medication side effects, how the person was feeling because of the TB disease, and social aspects that were mentioned. As a group, we will decide if this list covers all the important areas. If the group feels that

some are missing, these can then be added to the list of important areas relating to the patient's experience of TB disease and TB treatment. By the end of this focus group, we will have decided what we think are the most important areas that should be used to assess a new drug regimen for drug-resistant TB from the patient's perspective.

Once the list of important areas has been agreed the group will work to rank them. You have all been given a set of cards with the areas that seemed most important after the discussions in our first focus group, and we will now read through them together **[facilitator reads through all cards in view of participants and checks understanding]**.

<u>Intro (5-10 mins)</u>

- Remind participants that this stage of the discussion is about confirming that all points are of importance and making sure there are not any that have been missed. It is not yet about ordering the points
- *Does everyone agree that the points displayed on the cards are important for patients receiving treatment for drug-resistant TB?*
- *Are there any points that the group thinks are important, but are not presented here that could be added on the blank cards? In particular, are there any points relating to the effects from the disease itself that are not here?*
- If the majority agree on an item, the participants should add this item on to one of their blank cards in a manner that they find understandable (text, picture, or combination)
- The facilitator should ensure they read out the content added to the blank cards verbatim, for purposes of transcription

<u>Prioritising items for use in outcomes (10 – 15mins)</u>

- Provide each participant with fifteen small stickers
- Ask each person to place the stickers on the cards they consider most important from the set they have been provided. Participants can place as many of their available stickers on each card as they would like (e.g. on sticker on one card, two on another, and three on another to show levels of importance)
- The facilitator will collect all the cards back and count the total number of stickers each card has been awarded, and present the totals on a tally chart (presented on a flipchart, or other means of presenting to the whole group)
- The facilitator should ensure that the total number of stickers awarded to each item is both recorded in writing and read aloud at the time (for transcription purposes)

<u>Discussing the ordering of items (10 – 15mins):</u>

- *Does everyone feel comfortable with the items selected and the ordering according to the number of stickers?*
- The facilitator will then ask if the group can agree with assigning each item a high/medium/low importance status. The group will work through each item from highest-scoring to lowest-scoring and the facilitator will mark the item as "H/M/L"
- Items that received the same total number of stickers can be awarded different levels of importance after the group has discussed them

**Activity 3: Ordering of symptoms and side effects (20mins)**

- *We will now run a similar exercise to the last one, except looking at specific symptoms associated with TB disease and side effects from treatment. The aim of this next activity is to try to order these by how important the group thinks they are in terms of the patient's experience of having TB*

Ordering of Symptoms (10mins):

- Provide each participant with a set of 7 cards relating to TB symptoms and associated picture representation (cough, fever, weight loss, breathlessness, fatigue, and 2 blank) along with 10 small stickers
- The facilitator should read through all five cards using their own set displayed in a manner that allows all participants to see, and check understanding
- *Does everyone agree that these are the most important **symptoms** related to TB disease? Are there any that have not been included?*
- If the majority agree on an item, the participants should add this item on to one of their blank cards in a manner that they find understandable (text, picture, or combination)
- The facilitator should ensure they read out the content added to the blank cards verbatim, for purposes of transcription
- Ask each person to place the stickers on the cards they consider most important from the set they have been provided. Participants can place as many of their available stickers on each card as they would like (e.g. on sticker on one card, two on another, and three on another to show levels of importance)
- The facilitator will collect all the cards back and count the total number of stickers each card has been awarded, and present the totals on a tally chart (presented on a flipchart, or other means of presenting to the whole group)
- The facilitator should ensure that the total number of stickers awarded to each item is both recorded in writing and read aloud at the time (for transcription purposes)

Ordering of Side Effects (10mins):

- Provide each participant with a set of 7 cards relating to TB symptoms and associated picture representation (skin changes, nausea, diarrhoea, nerve problems, liver problems, and 2 blank) along with 10 small stickers
- The facilitator should read through all five cards using their own set displayed in a manner that allows all participants to see, and check understanding
- *Does everyone agree that these are the most important **side effects** related to TB treatment? Are there any that have not been included?*
- If the majority agree on an item, the participants should add this item on to one of their blank cards in a manner that they find understandable (text, picture, or combination)
- The facilitator should ensure they read out the content added to the blank cards verbatim, for purposes of transcription
- Ask each person to place the stickers on the cards they consider most important from the set they have been provided. Participants can place as many of their available stickers on each card as they would like (e.g. on sticker on one card, two on another, and three on another to show levels of importance)
- The facilitator will collect all the cards back and count the total number of stickers each card has been awarded, and present the totals on a tally chart (presented on a flipchart, or other means of presenting to the whole group)
- The facilitator should ensure that the total number of stickers awarded to each item is both recorded in writing and read aloud at the time (for transcription purposes)

**Debrief to close session (5mins):**

- Thank everyone for participating in the focus group and for contributing valuable insights that will help with the development of better tools for researching new drugs to help fight TB
- Ask the group if they feel there is anything that has not been covered in the focus group that should have been

- Let the group know that they should approach the facilitators after the session if there were any parts of the discussion that they found distressing or difficult for any reason
- Finally, ensure the group knows that any feedback on how the group discussions could have improved is welcome and they can share this with the group now or approach a facilitator afterwards
